# *In vivo* Assessment of Brain Microstructure in Patients with Huntington’s Disease Using Quantitative MRI

**DOI:** 10.64898/2026.01.18.26344351

**Authors:** Marta M. Pokotylo, Joke-Lina Aßmann, Maximilian G. Ködderitzsch Mertins, Julia Henkel, Jan Uter, Leon van Well, Alexander Münchau, Sebastian Löns, Martin Gottlich, Norbert Brüggemann, Jannik Prasuhn

## Abstract

**Objectives:** Utilizing the quantitative magnetic resonance imaging (MRI) technique, multiparametric mapping (MPM), we assessed the integrity of cortical and subcortical regions across different Huntington’s disease (HD) stages, relating the derived findings to genetic modifiers and clinical measures.

**Methods:** Sixteen premanifest patients with HD (PwHD_pre_), ten manifesting PwHD (PwHD_man_), and forty-two healthy controls (HC) underwent clinical examinations and an MRI session at 3T. We derived quantitative MPM maps and performed voxel-based quantification (VBQ) and voxel-based morphometry (VBM). We assessed the group differences between the combination of PwHD_pre_ and PwHD_man_ (PwHD_total_), PwHD_pre_ and PwHD_man_, compared with matched HC. Using correlation analysis, we investigated the link between MPM-derived findings, CAG repeat length, and clinical scores.

**Results:** PwHD_man_ presented with reductions in all four quantitative MPM maps across bilateral caudate nucleus, frontal, parietal, temporal, and occipital gyri. We observed spatial overlap between VBQ- and VBM-derived findings, demonstrating widespread volume loss in PwHD_man_. PwHD_pre_ presented with reductions in some quantitative maps within frontal and temporal regions. Reduced MPM values correlated with clinical scores of HD severity.

**Interpretation:** The observed reductions across four MPM modalities in PwHD_man_ are consistent with demyelination and neuronal loss, supported by observations of widespread volume loss. While the observed reductions in PwHD_pre_ are indicative of early demyelination and inflammation. In this study, we demonstrated the utility and clinical relevance of MPM for detecting microstructural alterations across different stages of HD. MPM can serve as a non-invasive biomarker of neurodegeneration in HD, with insights into pathological events underlying the observed neuronal loss.

## INTRODUCTION

Huntington’s disease (HD) is an autosomal-dominant neurodegenerative disease caused by an expanded CAG repeat in the *huntingtin (HTT)* gene, with a greater repeat length inversely correlating with onset and greater disease severity(1,2). Although the characterization of HD causes, there are no approved disease-modifying treatments yet, with only symptomatic strategies available(3). Furthermore, patients with HD (PwHD) exhibit highly heterogeneous symptomatology, spanning across motor, cognitive, and psychiatric domains, with emerging studies demonstrating symptom worsening with disease progression. Additionally, PwHD subgroups experience a more pronounced decline in specific domains, for reasons not yet understood(4,5). Such heterogeneous clinical presentation and disease progression, along with variable responses to symptomatic treatment, demonstrated the need for improved biomarkers that can improve patient stratification and monitoring.

Magnetic resonance imaging (MRI) is the most utilized neuroimaging tool for the detection of the basal ganglia (BG) atrophy, one of the hallmarks of HD pathology, correlating with disease motor symptom severity and disease duration(5,6). However, MRI is limited to the detection of macroscopic structural changes, falling short in probing underlying molecular and pathogenic mechanisms(7). Quantitative MRI (qMRI), such as multiparametric mapping (MPM), offers an alternative with higher sensitivity, assessing qualitative measures of tissue’s biophysical properties, allowing the assessment of brain microstructural and cellular level composition(8). By modeling signals from multi-contrast gradient-echo acquisitions, MPM quantifies longitudinal relaxation rate (R1), effective transverse relaxation rate (R2*), proton density (PD), and magnetic transfer saturation (MTsat)(9,10). As R1 and MTsat are sensitive to myelin, R2* to iron, and PD to water content, MPM allows *in vivo* assessment of these tissue characteristics and underlying pathogenic processes, deriving insights previously obtainable only with histology. As histology techniques assess the end-stage of brain microstructure, their clinical relevance is limited, making MPM a better clinical alternative(8).

Despite its strong, clinically relevant potential, the use of MPM in HD research and clinical practice has not been explored. Therefore, we utilized MPM to assess the cortical, white matter (WM), and BG microstructural differences between manifesting PwHD (PwHD_m_an), premanifest PwHD (PwHD_pre_), and healthy controls (HC). Given the evidence of widespread cortical neurodegeneration in HD, we hypothesized that MPM insights would reveal widespread cortical and BG atrophy and demyelination, associated with the clinical measures of HD severity(11,12).

## METHODS

### Experimental design and participants

*We* recruited participants from inpatient and outpatient clinics at the Center for Rare Diseases of the University-Medical Center Schleswig-Holstein (Campus Lübeck). Participants aged 50-80 years without concomitant neurological disorders were included. PwHD were selected based on a genetically confirmed diagnosis with fully-penetrant, ≥ 40 CAG repeats in the *HTT* gene(13). Exclusion criteria included MRI contraindications, comorbidities affecting informed consent, or a history of structural brain diseases. All PwHD underwent clinical examinations, including the Unified Huntington’s Disease Rating Scale (UHDRS) and cognitive screenings with the Montreal Cognitive Assessment (MoCA)(14,15). The study adhered to the Declaration of Helsinki and was approved by the local ethics committee (University of Lübeck, AZ-18-196).

### Image acguisition, voxel-based quantification, and voxel-based morphometry

MRI sessions were conducted using a 3T Siemens Magnetom Skyra scanner equipped with a 64-channel head coil. Using a multi-echo 3D fast low-angle shot, we acquired T1, PD, and magnetization transfer (MT) weightings(16). We measured all sequences in coronal orientation with 240 contiguous slices, 1.0 mm isotropic resolution, a field of view of 256 × 176 mm^2^ (phase FoV 68.8 %), GRAPPA acceleration factor 2 with 24 reference lines, and partial Fourier 6/8 in the slice direction. The received bandwidth for all echoes was 440 Hz/p×. We enabled inline distortion correction (2D mode) and prescan normalization. The T1-weighted acquisition (TR = 19.0 ms, flip angle = 20°, six echoes with TE = 2.20/4.70/7.20/9.70/12.20/15.00 ms) required about 8 minutes. The PD-weighted acquisition (TR = 24.0 ms, flip angle = 6°, eight echoes with TE = 2.20/4.70/7.20/9.70/12.20/15.00/17.70/20.00 ms) required approximately 7 minutes. The magnetic transfer-weighted acquisition (TR = 37.0 ms, flip angle = 6°, six echoes with TE = 2.20/4.70/7.20/9.70/12.20/15.00 ms, with an off-resonance magnetization transfer pre-pulse enabled) required about 11 minutes. The neuroimaging data were reviewed by a neuroradiologist to exclude concurrent conditions. We performed a visual assessment of artifacts (e.g., head motion) and quantitative evaluations comprising calculating the number of artifact voxels (an exclusion threshold was set to artifacts exceeding two standard deviations above the mean).

Voxel-based quantification (VBQ) analysis was performed in hMRI toolbox within the Statistical Parametric Mapping 25 framework (SPM25, Wellcome Trust Center for Neuroimaging, London) in MATLAB software (R2025a, The MathWorks Inc., Natick, MA, USA)(8). We reconfigured the toolbox with a custom-written, scanner-specific settings MATLAB script. We created and processed R1, R2*, PD, and MTsat maps from unprocessed, multi-echo T1-, PD-, and MT-weighted images, using the ‘UNICORT’ setting for B1 and unified segmentation for radiofrequency sensitivity bias corrections(8). We used the hMRI toolbox pipeline for processing, comprising segmentation (into gray matter (GM) and WM); nonlinear spatial registration to MNI space; and tissue-weighted smoothing(17). We set a voxel size of 1 × 1 × 1 × mm^3^, a bounding box of 2 × 3, and Gaussian smoothing with a 6 × 6 × 6 mm FWHM kernel.

For voxel-based morphometry (VBM) analysis, we utilized the Computational Anatomy Toolbox 12 (CAT12.9) and R1 maps generated during VBQ analysis(18).

### Statistical analyses

Using SPM25, we conducted VBQ and VBM second-level analysis involving a two-sample t-test. Age and sex were included as covariates for VBQ, and additionally, total intracranial volume for VBM analyses. The cluster-forming threshold was set to p < .001 (uncorrected), with a minimum number of voxels of 20; additional cluster-level family-wise error (FWE) correction of p < .05 was applied. We tested two contrasts: clusters with higher values in PwHD (PwHD > HC) and clusters with lower values in PwHD (PwHD < HC). Using MarSBaR (v0.45), we extracted clusters with significant group-level differences and visualized them using Python software (v3.13.5, Python Software Foundation) and libraries such as Nilearn (v.0.12.0)(19,20).

We performed two sets of group comparisons. The two-group comparison comprised all PwHD participants, PwHD_total_ (PwHD_pre_ and PwHD_man_), versus age- and sex-matched HC_total_. The four-group comparison comprised PwHD_pre_ versus HC_pre_; and PwHD_man_ versus HC_man_. We statistically stratified HC_pre_ and HC_m_an from the HC_total_ group. HC groups comprise exclusively healthy participants; “pre” and “man” labeling refers only to the matched PwHD group.

We performed separate analyses for whole-brain and the BG; VBQ and VBM; each MPM map; as well as for GM and WM, using explicit masks. We used tissue probability masks incorporated within the SPM25 environment, and the BG mask was created using the ATAG atlas(21). Significant clusters were anatomically labeled using the Neuromorphometrics atlas for GM and the JHU White-Matter Tractography Atlas for WM(22).

Using Pearson’s and Spearman’s correlation tests, we explored relationships between mean MPM values in significant clusters and clinical measures, including UHDRS, MoCA, disease duration, and the CAG repeat length *(mHTT).* Using MarSBaR, we extracted mean parameter values within the cluster of interest, using them as predictors in correlative analysis. For ross-modal correlation analysis, we used clusters demonstrating significant group differences that survived Bonferroni correction across modalities (four MPM maps × two contrasts, p < .00625).

## RESULTS

### Demographic and Clinical Assessment

*We* included 26 PwHD_total_ (age: 47.1 ± 14.9; 15 females) and 42 HC_total_ (age: 49.0 ± 18.0; 22 females). The mean age (p = 0.649) or sex distribution (χ^2^ = .000, df = 1, p = .525) did not differ between groups. Within the PwHD_total_ group, 16 were PwHD_pre_ (age: 39.2 ± 12.4; 10 females), and 10 were PwHD_man_ (60.9 ± 7.1; 6 females), with no age or sex differences relative to their corresponding HC groups (PwHD_pre_: age: p = .938; sex distribution: χ^2^ = .000, df = 1, p = 1.000. PwHD_man_: age: p = .757; sex distribution χ^2^ = .000, df = 1, p = 1.000). The average CAG repeat length was 42 ±2 among PwHD_pre_, and 41 ± 1 among PwHD_man_. Mean disease duration and UHDRS scores in PwHD_man_ were 7.7 ± 5.5 years and 42.40 ± 19.72, respectively. The average MoCA scores were 26.56 ± 2.59 in the PwHD_pre_ group and 19.70 ± 5.38 in the PwHD_man_ group (Table 1).

**Table 1.** Demographic and clinical characteristics.

| Two-group comparison |  |  |  |  |  |  |
| --- | --- | --- | --- | --- | --- | --- |
|  | PwHD<sub>total</sub> | HC<sub>total</sub> | p-value |  |  |  |
| number | 26 | 42 | .649<sup>1</sup> |  |  |  |
| male/female | 11/15 | 20/22 | .525<sup>2</sup> |  |  |  |
| age (years) | 47.1 ± 14.9<br>[35.2 – 60.5] | 49.0 ± 18.0<br>[30.7 – 63.0] |  |  |  |  |
| mHTT<br>(no. of CAG<br>repeats) | 42 ± 2<br>[40 – 43] | n.a. |  |  |  |  |
| UHDRS | 17.50 ± 23.35<br>[1.25 – 23.75] | n.a. |  |  |  |  |
| Disease<br>duration (years) | 7.7 ± 5.5<br>[5.2 – 9.2] | n.a. |  |  |  |  |
| MoCA | 23.92 ± 5.10<br>[22.00 – 28.00] | n.a |  |  |  |  |
| Four-group comparison |  |  |  |  |  |  |
|  | PwHD<sub>pre</sub> | HC<sub>pre</sub> | p-value | PwHD<sub>man</sub> | HC<sub>man</sub> | p-value |
| number | 16 | 16 |  | 10 | 10 |  |
| male/female | 6/10 | 6/10 | 1.000<sup>1</sup> | 4/6 | 4/6 | 1.000<sup>1</sup> |
| age (years) | 39.2 ± 12.3<br>[31.8 – 43.5] | 39.3 ± 14.5<br>[28.0 – 52.0] | .938<sup>2</sup> | 59.8 ± 8.5<br>[53.0 – 65.7] | 60.9 ± 7.1<br>[56.7 – 63.0] | .757<sup>2</sup> |
| mHTT<br>(no. of CAG<br>repeats) | 42 ± 2<br>[41 – 43] | n.a. |  | 41 ± 1<br>[40 – 43] | n.a. |  |
| UHDRS | 1.94 ± 1.98<br>[0.00 – 2.25] | n.a. |  | 42.40 ± 19.72<br>[23.25 – 57.25] | n.a. |  |
| Disease<br>duration (years) | n.a. | n.a. |  | 7.7 ± 5.5<br>[5.2 – 9.2] | n.a |  |
| MoCA | 26.56 ± 2.59<br>[24.25 – 29.00] | n.a |  | 19.70 ± 5.38<br>[17.25 – 24.50] | n.a |  |

Mean values and standard deviations are quoted. Values in square brackets represent the interquartile range. <sup>1</sup> according to a $\chi^2$ -test; <sup>2</sup> two-sample t-test applied. HC<sub>pre</sub> and HC<sub>man</sub> groups comprise exclusively healthy participants. The “pre” and “man” labels are used to facilitate clarity when reading our work. HC<sub>pre</sub>/HC<sub>man</sub>: healthy controls. mHTT: number of CAG repeats in the expanded *Huntington* gene. MoCA: Montreal Cognitive Assessment. n.a.: not applicable. PwHD<sub>man</sub>: manifesting patients with Huntington’s disease. PwHD<sub>pre</sub>: premanifest patients with Parkinson’s disease. PwHD<sub>total</sub>: premanifest and manifesting patients with Huntington’s disease. UHDRS: Unified Huntington’s Disease Rating Scale.

### Widespread microstructural cortical and subcortical reductions in patients with Huntington’s disease

We observed constant reductions in t-values across all four MPM modalities in PwHD_total_ (Figure 1, Supplementary Tables 1-4). Identified clusters included bilateral middle temporal gyri (left MTG: R1: T = 5.43, p_FWE_ = .000; R2*: T = 3.93, p_FWE_ = .005; PD: T = 5.64, p_FWE_ = .000. Right MTG: R1: T = 5.33, p_FWE_ = −001; MTsat: T = 4.83, p_FWE_ = .000), superior temporal gyri (left STG: R2*: T = 4.99; MTsat: T = 6.00; right STG: R2*: T = 4.82; MTsat: T = 4.52; all p_FWE_s = .000), left posterior orbital gyrus (POrG: R1: T = 3.87, p_FWE_ = .000; R2*: T = 4.16, p_FWE_ = .000: MTsat: T = 4.38, p_FWE_= .002), left fusiform gyrus (FuG: R1: T = 6.68, p_FWE_ = .041; R2*: T = 6.62, p_FWE_ = .010; MTsat: T = 6.48, p_FWE_ = .023), left supramarginal gyrus (SMG: R2*: T = 5.19, p_FWE_ =-000; MTsat: T = 4.76, p_FWE_ = .021) and left postcentral gyrus (PoG: R1: T = 4.14, p_FWE_ = .000, R2*: T= 5.78, p_FWE_ = .000; MTsat: T = 4.28, p_FWE_ = .001). Additionally, reductions in PD were observed in bilateral caudate nuclei (left Caudate: T = 4.77; right Caudate: T = 4.50; all p_FWE_s = .000). VBM analysis revealed only clusters with lower t-values in PwHD_total_, including left Caudate (T = 5.60, p_FWE_ = .000) and right middle frontal gyrus (T = 4.99, p_FWE_ = .024), which overlapped with VBQ,-identified clusters (Supplementary Table 5).

Within BG, t-values were lower in PwHD_total_ across all MPM modalities in bilateral Caudate (left: R1: T = 3.63, p_FWE_ = .041; R2*: T = 3.86, p_FWE_ = .017; MTsat: T = 4.33, p_FWE_ = .000. Right: R1: T = 4.07, p_FWE_ = .000; R2*: T = 4.40, p_FWE_ = .000; PD: T = 3.86, p_FWE_ = .040; MTsat: T = 4.14, p_FWE_ = .000, Supplementary Figures 1, 2, and Table 6). VBM-derived clusters with lower t-values were identified in bilateral Caudate (left: T = 5.27; right: T = 4.59; all p_FWE_s = .000).

### Altered R2*, RI, and MTsat in premanifest patients with Huntington’s disease

PwHD_pre_ presented with differences across R1, R2*, and MTsat only (Figure 2; Supplementary Table 7), including lower R1 values in the left superior frontal gyrus (SFG, T = 5.53, p_FWE_ = .028) and left MTG (T = 3.95, p_FWE_ = .031); lower R2* values (all p_FWE_s = .000) in the right middle frontal gyrus (MFG, T = 5.26) and right superior parietal lobule (SPL, T = 4.84); and lower MTsat values in right SFG (T = 5.82, p_FWE_ = −001) and right posterior cingulate gyrus (PCgC, T = 4.45, p_FWE_ = −014) in PwHD_pre_ (Figure 2). No significant group differences were observed in whole-brain VBM; BG VBQ and VBM analyses.

### Multimodal reductions in cortical and subcortical structures in manifesting patients with Huntington’s disease

All identified clusters presented with lower t-values in PwHD_man_ (Figure 3; Supplementary Tables 8-11). The left STG presented with consistent reduction across all MPM modalities (R1: T = 10.65; R2*: T = 5.09; all p_FWE_s = .000; PD: T = 6.96; MTsat: T = 10.72, all p_FWE_s = .016). Multiple clusters demonstrated reduced R1 and R2* values, including the bilateral postcentral gyrus (left PoG: R1: T = 8.21, p_FWE_ = −000; R2*: T = 6.67, p_FWE_ = −002. Right: R1: T = 6.71, p_FWE_ = .028; R2*: T = 6.60, p_FWE_ = .000), the left middle cingulate gyrus (MCgG: R1 = 8.20, p_FWE_ = −003; R2*: T = 8.10, p_FWE_ = .002), and the left inferior occipital gyrus (IOG: R1 = 4.77; R2*: T = 9.14, all p_FWE_S = .000). Lower MTsat (all p_FWE_S = .000) values were detected in cerebellar structures, including left cerebellum exterior (T = 4.65) and vermal lobules VI l-VI 11 (T = 7.16). All VBM-identified clusters overlapped with VBQ-derived clusters, including within the frontal (right SFG: T = 7.33, p_FWE_ = −001; right MFG: T = 6.45, p_FWE_ = −023; and left MFG: T = 6.42, p_FWE_ = .000) and parietal (right SPL: T = 7.33, p_FWE_ = .023; and right PoG: T = 10.46, p_FWE_ =.000) lobes (Supplementary Table 12). Furthermore, the largest VBM-identified clusters in the right FuG (T = 16.32, p_FWE_ = .000) overlapped with lower R2* values (T = 5.51, p_FWE_ = .033).

The BG analysis demonstrated reduced R2* and MTsat values in PwHD_man_ only (Supplementary Figure 3, Table 13). Right Caudate presented with lower R2* (T = 4.75, p_FWE_ = .001) and MTsat values (T = 5.23, p_FWE_ = .000), while left Caudate presented with reduced MTsat values only (T = 6.06, p_FWE_ = .000). All VBQ-derived clusters overlapped with VBM-derived reductions (all p_FWE_S = .000) in right (T = 5.37) and left putamen (T = 5.07).

### Overlapping and correlated cross-modal group differences within the same clusters

In PwHD_total_, reductions across MPM modalities overlapped within left FuG, left MTG, right thalamus, and left middle occipital gyrus (MOG) (Figure 4, Table 2). Lower R1 values positively correlated with lower R2* values (all ps < .0001) in left FuG (r = 0.953), right thalamus (r = 0.955), bilateral Caudate (left: r = 0.690; right: r = 0.694), and left MOG (r = 0.551, p = .0035); and with lower MTsat (all p < .0001) in left FuG (r = 0.836), right thalamus (r = 0.707), and bilateral Caudate (left: r = 0.190; right r = 0.934). We identified positive correlations (all ps < .0001) between lower R1 and PD in left MTG (r = 0.966), and lower R2* and MTsat in left FuG (r = 0.892), right thalamus (r = 0.691), and bilateral Caudate (left: r = 0.753; right: r = 0.837). Similarly, in PwHD_man_, RI and R2* values positively correlated in left STG (r = 0.660, p = .0438), left PoG (r = 0.878, p = .0016), left MCgG (r = 0.987, p < .0001), right PoG (r = 0.927, p = .0003), and left IOG (r = 0.745, p = .0174). Additionally, in ISTG, RI positively correlated with MTsat values (r = 0.672, p = .0390) in PwHD_man_.

**Table 2.** Cross-modal correlations between MPM modalities in gray matter clusters with significant group-level differences.

| Anatomical location | Affected modalities |  | r | p-value |
| --- | --- | --- | --- | --- |
| PwHD<sub>total</sub> vs HC<sub>total</sub> |  |  |  |  |
| lFuG | R1 | R2* | .953<sup>*</sup> | p < .0001 |
|  | R1 | MTsat | .836<sup>*</sup> |  |
|  | R2* | MTsat | .892 |  |
| lMTG | R1 | PD | .966<sup>*</sup> | p < .0001 |
| rThal | R1 | R2* | .955<sup>*</sup> | p < .0001 |
|  | R1 | MTsat | .707<sup>*</sup> |  |
|  | R2* | MTsat | .691<sup>*</sup> |  |
| lMOG | R1 | R2* | .551<sup>*</sup> | p = .0035 |
| lCaudate | R1 | R2* | .690<sup>*</sup> | p < .0001 |
|  | R1 | MTsat | .910<sup>*</sup> |  |
|  | R2* | MTsat | .753<sup>*</sup> |  |
| rCaudate | R1 | R2* | .694<sup>*</sup> | p < .0001 |
|  | R1 | MTsat | .934<sup>*</sup> |  |
|  | R2* | MTsat | .837<sup>*</sup> |  |
| PwHD<sub>man</sub> vs HC<sub>man</sub> |  |  |  |  |
| lSTG | R1 | R2* | .660<sup>*</sup> | p = .0438 |
|  | R1 | MTsat | .672<sup>*</sup> | p = .0390 |
| lPoG | R1 | R2* | .878<sup>*</sup> | p = .0016 |
| lMCgG | R1 | R2* | .987<sup>*</sup> | p < .0001 |
| rPoG | R1 | R2* | .927<sup>*</sup> | p = .0003 |
| lIOG | R1 | R2* | .745<sup>*</sup> | p = .0174 |
| rCaudate | R2* | MTsat | .699 | p = .0243 |

\* according to Spearman's correlation. lCaudate: left caudate nucleus. lFuG: left fusiform gyrus. lIOG: left inferior occipital gyrus. lMCgG: left middle cingulate gyrus. lMOG: left middle occipital gyrus. lMTG: left middle temporal gyrus. lPoG: left postcentral gyrus. lSTG: left superior temporal gyrus. The HC<sub>man</sub> group comprises only healthy participants, and “man” labeling refers to the matched PwHD<sub>man</sub> group. HC<sub>total/man</sub>: healthy controls. MPM: multiparametric mapping. MTsat: magnetic transfer saturation. PD: proton density. PwHD<sub>man</sub>: manifesting patients with Huntington's disease. PwHD<sub>total</sub>: premanifest and manifesting patients with Huntington's disease. r: correlation coefficient. R1: longitudinal relaxation rate. R2\*: effective transverse relaxation rate. rCaudate: right caudate nucleus. rPoG: right postcentral gyrus. rThal: right thalamus.

### Identified group differences across MPM modalities were associated with clinical variables

*We* detected significant associations between MPM-derived values and clinical parameters in PwHD_total_ (Figure 5; Supplementary Figure 4). Lower R1, R2*, PD, and MTsat values in the bilateral Caudate negatively correlated with UHDRS and disease duration and positively correlated with MoCA scores. Lower R2* in the right MOG (r = 0.927, p = .0001) and left precuneus (r = 0.884, p = .0069) correlated with higher MoCA scores. Only few clusters demonstrated positive correlations with the CAG repeat length, including left MCgG R1 (r = 0.636, p = .0483) and R2* values (r = 0.636, p = .0483), R2* values in the left SMG (r = 0.397, p = .0387) and right precentral gyrus (r = 0.653, p = .0372); and lower MTsat within left STG (r = 0.685, p = .0287).

### VBQ- and VBM-derived white matter microstructural and volumetric changes

We observed multimodal reductions in both PwHD_total_ and PwHD_pre_; however, no VBM differences were detected in these groups (Supplementary Figures 5, 6, Table 14-15). We detected no VBM differences in PwHD_total_ or PwHD_pre_.

Within the PwHD_man_ group, we detected MPM reductions (all p_FWE_s = .000), within the splenium (sCC: R1: T = 21.58; R2*: T = 57.4; MTsat: T = 5.09) and genu of the corpus callosum (gCC: R1: T = 6.56; R2*: T = 6.16; MTsat: T = 5.83) (Supplementary Figure 7, Table 16). Consistent with GM findings, MPM-derived clusters overlapped with VBM-derived reductions in t-values in PwHD_man_ in posterior corona radiata (left: T = 7.52, p_FWE_ = .011; right: T = 5.65, p_FWE_ = .015), right superior corona radiata (T = 6.16, p_FWE_ = .016), sagittal stratum (T = 6.26, p_FWE_ = .000), and left cingulum (T = 6.11, p_FWE_ = .000, Supplementary Table 17). Some of the clusters with identified VBQ reductions cross-modally overlapped (Supplementary Figure 8).

## DISCUSSION

Utilizing MPM, we detected microstructural alterations in PwHD. PwHD_total_ presented with widespread cortical and subcortical reductions across all MPM parameters, while in PwHD_man_, we observed more spatially constrained changes with stronger group effects.

Reductions in R1 and MTsat are reflective of myelin and macromolecular content loss. Reduced R2* reflects reduced iron content and demyelination, while reduced PD represents a reduction in water content, aligning with tissue loss(23,24). The extensive cross-modal overlap of MPM-derived findings is consistent with progressive neurodegeneration and demyelination, supported by spatial overlap with VBM-derived findings of widespread cortical and subcortical GM and WM volume loss.

Bilateral caudate nuclei consistently demonstrated reductions across all MPM maps in PwHD_man_, indicative of neurodegeneration, while reductions in R2* and MTsat in PwHD_pre_ suggest that these markers capture ongoing pathology more robustly. Findings of negative correlations between MPM-based differences and UHDRS, disease duration, and a positive correlation with MoCA scores are consistent with available literature, emphasizing the clinical relevance of MPM in HD research(25). Within the BG, no MPM- or VBM-derived differences were observed in PwHD_pre_, despite evidence of BG atrophy preceding clinical onset, possibly reflecting limited sensitivity to subtle microstructural changes or limited statistical power (n = 16)(26). Alternatively, BG is functionally segregated into motor, associative, and limbic circuits, requiring further analyses with the utilization of functional BG atlases to provide deeper insights with higher sensitivity into early microstructural changes(27). Although studies utilizing quantitative susceptibility-weighted mapping (QSM) demonstrated iron accumulation within the BG in PwHD, we demonstrated reduced R2*, reflecting reduced iron content(28). As QSM is sensitive to iron and myelin, high QSM values reflect iron accumulation and demyelination; findings are not directly comparable to R2*(29,30).

Spatially restricted clusters with higher R2* and MTsat in PwHD_pre_ reflect local iron accumulation and neuroinflammation, demonstrating early-stage alterations with regional susceptibility. Findings of MPM-derived alterations in MFG, SFG, and MTG and VBM-derived volume loss in the right middle orbital gyrus in PwHD_pre_, are consistent with widespread neuronal loss prior to disease clinical onset(11,12).

The two- and four-group comparisons revealed distinct spatial patterns in MPM-related alterations in PwHD_total_ and PwHD_man_, with PwHD_total_ demonstrating the core disease-related microstructural alterations, while the PwHD_man_ presented spatially confined clusters with stronger effect sizes, reflecting disease-stage vulnerability. We observed consistent reductions across all four MPM modalities in bilateral caudate nucleus, thalamus, MFG, STG, postcentral gyri, and occipital gyri, aligning with available evidence, adding insights into underlying biophysical changes and mechanisms driving the observed neurodegeneration(31,32). Furthermore, our findings are supported by longitudinal studies and postmortem studies demonstrating progressive neurodegeneration with disease progression(33–35).

Although MPM alterations in PwHD_man_ were associated with clinical measures, no single modality could be attributed to a specific clinical measure. Given the multisystem and progressive nature of the HD, the identified changes are best explained as contributors rather than sole causes of clinical presentation(36–38).

The similar pattern of reductions across MPM modalities in WM demonstrates widespread microstructural alterations across both types of tissue. Although WM degeneration has been demonstrated to precede GM loss, we did not replicate this, detecting no volumetric loss in PwHD_pre_(39). Given that diffusion tensor imaging (DTI) is the most utilized technique for the assessment of WM integrity, relying on fundamentally different physical principles than MPM, it is challenging to cross-compare the findings. However, progressive reduction of R2* from PwHD_pre_ to PwHD_man_, demonstrates that early myelin changes occur before neuronal loss in later stages of HD.

Our study has several limitations, including the cross-sectional design. We aimed to demonstrate the MPM’s utility in HD research, and future studies should include longitudinal trajectories to assess the disease stage-dependent progression of MPM-derived findings. Additionally, disease stage stratification into PwHD_pre_ and PwHD_man_ cohorts reduced the statistical power and the ability to detect more subtle microstructural changes, specifically in PwHD_pre_. The narrow range of CAG repeat lengths within our cohort may limit our ability to detect significant correlations between repeat length and MPM-derived metrics. Therefore, future studies should aim to have larger and more heterogeneous cohorts of PwHD. The used measure of disease severity comes with its own limitations, including intra- and inter-rated variability and subjective bias, while MoCA tests assess the general state of cognition with no domain-specific insights(40—42). Further studies may use multi-domain composite UHDRS and a more representative panel of cognitive and neuropsychological assessments, as well as integration of genetic expression profiling and post-mortem investigation, to get greater insights into the association between MPM-derived findings and specific domains of symptomatology and underlying cellular compositions(38,43).

To conclude, utilizing MPM, we identified widespread cortical and subcortical microstructural alterations across HD stages, with consistent reductions consistent with neurodegeneration. These observations were spatially overlapping with volumetric reductions and were associated with clinical measures in PwHD, supporting the utility of MPM for *in vivo* assessment of disease-related pathology in HD. Given the high insights output within a single scan, MPM has great potential for improving diagnostics and patient stratification, as well as personalized treatment approaches, necessitating confirmation in future longitudinal studies.

## Data Availability

All data produced in the present study are available upon reasonable request to the authors

## ACKNOWLEDGMENT

We would like to acknowledge Jenny Schmalfeld for her assistance in participant recruitment and study coordination.

## AUTHORS CONTRIBUTION

**MMP:** Conceptualization, Methodology, Data curation, Formal analysis, Visualization, Writing the original draft. JLA: Investigation, Data curation, Writing review and editing. **MGKM:** Investigation, Data curation, Writing review and editing. **JH:** Investigation, Data curation, Writing review and editing. JU: Investigation, Data curation, Writing review and editing. **LvW:** Investigation, Data curation, Writing review and editing. **AM:** Resources, Clinical expertise, Writing review and editing. SL: Investigation, Data curation, Writing review and editing. NB: Supervision, Project administration, Funding acquisition, Writing review and editing. JP: Conceptualization, Methodology, Supervision, Writing review and editing.

## STATEMENT CONFIRMING AVAILABILITY OR ABSENCE OF SHARED DATA

The data generated in this study is not publicly available. De-identified data may be provided by the corresponding authors upon reasonable request, subject to institutional approval.

---

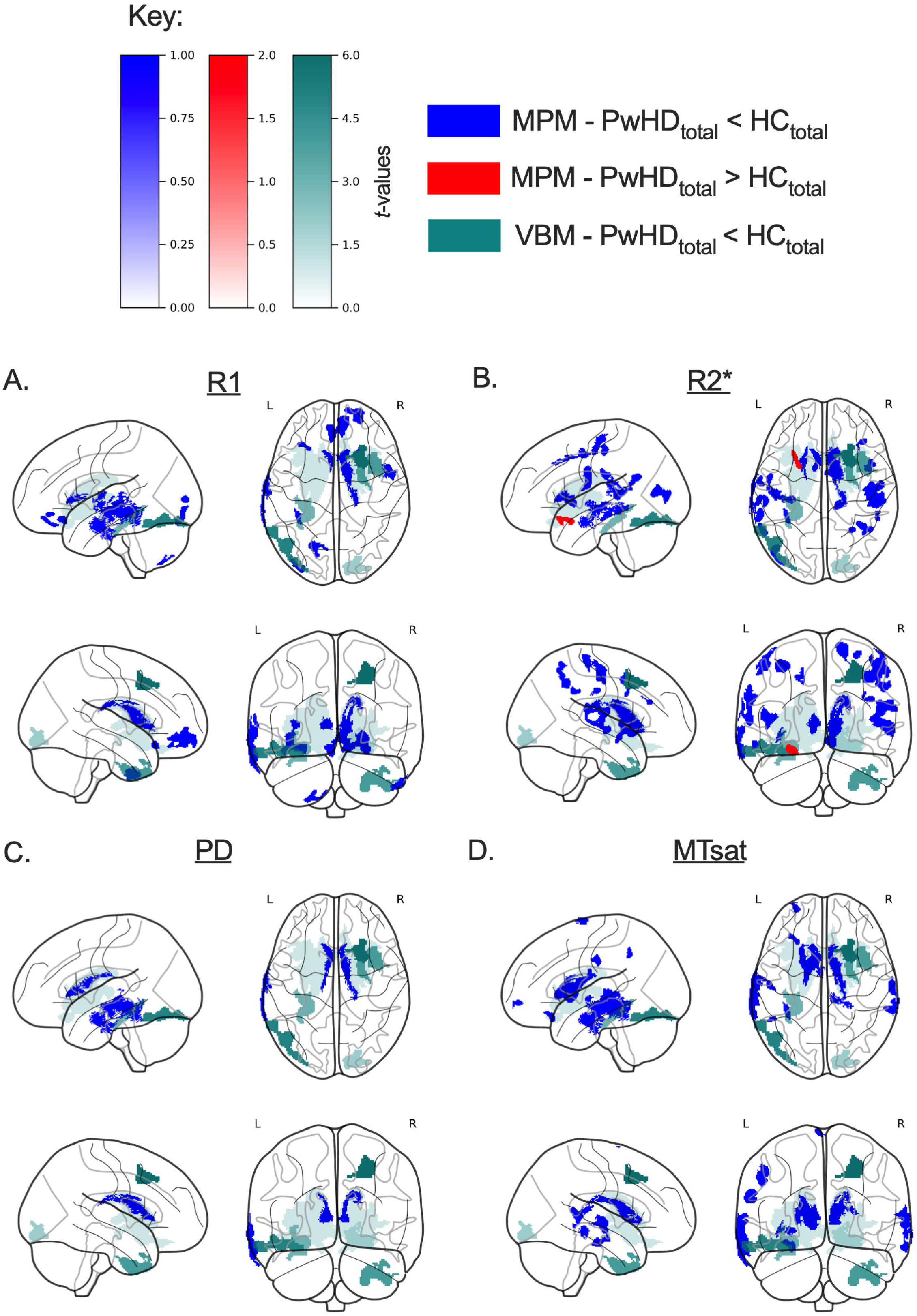

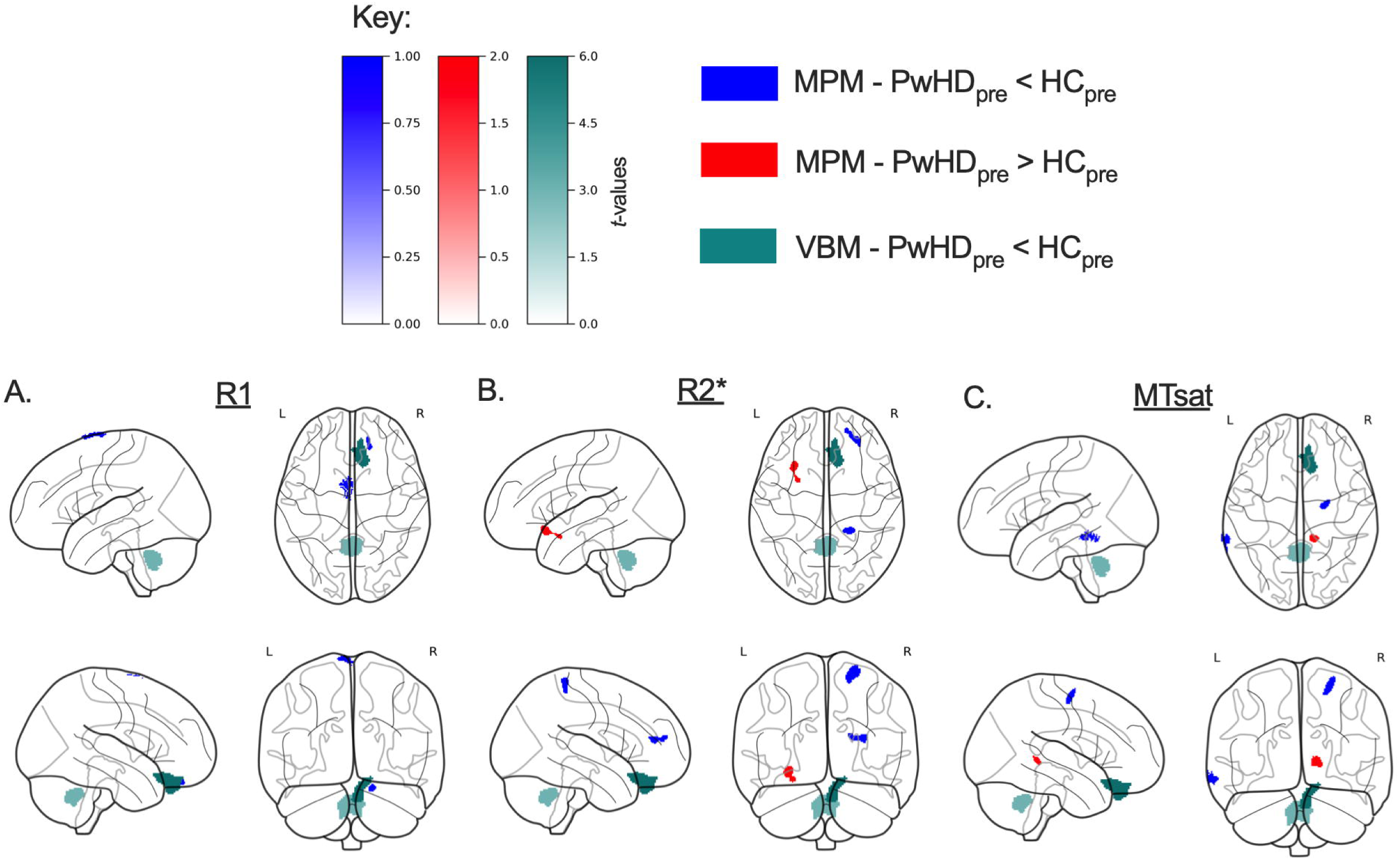

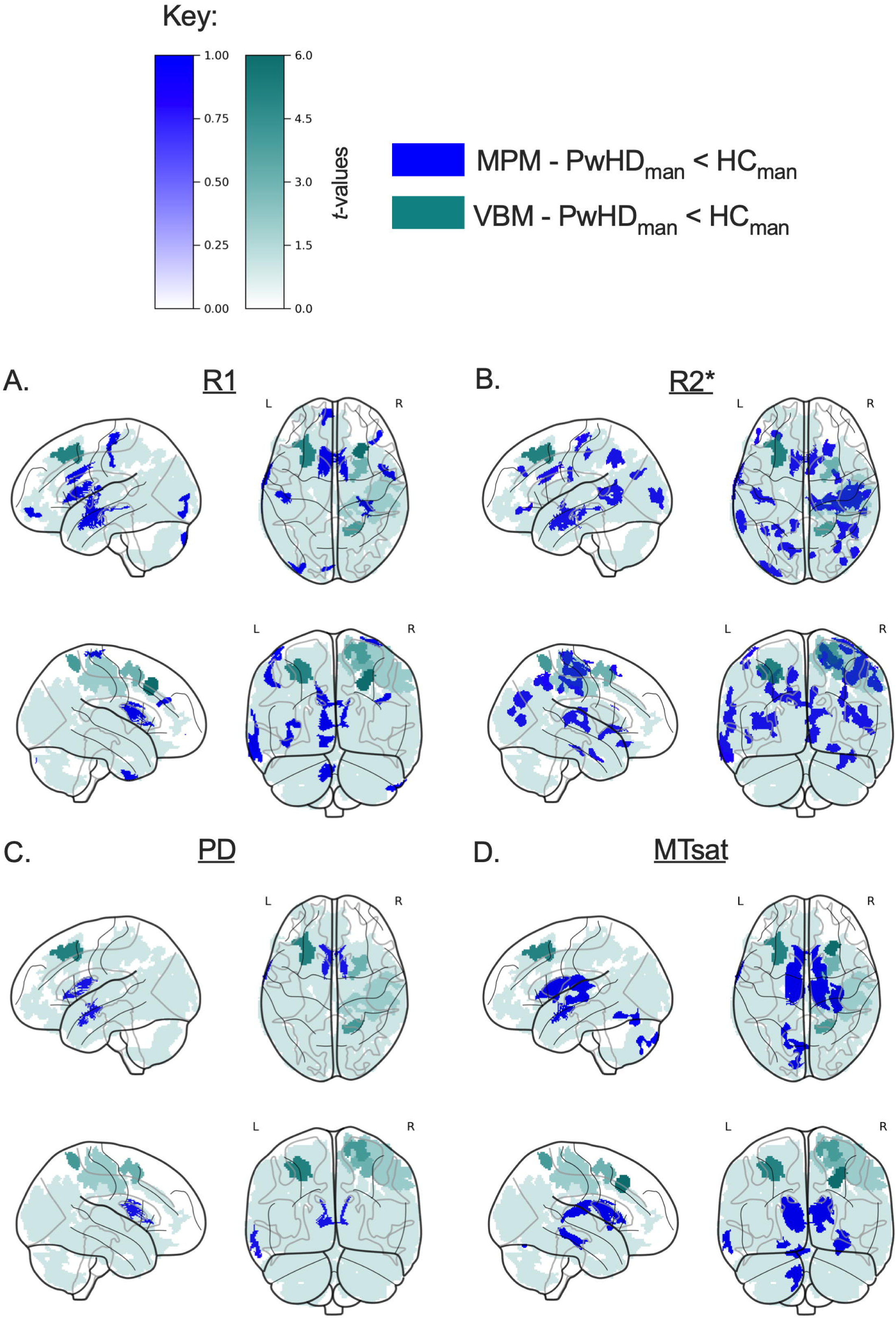

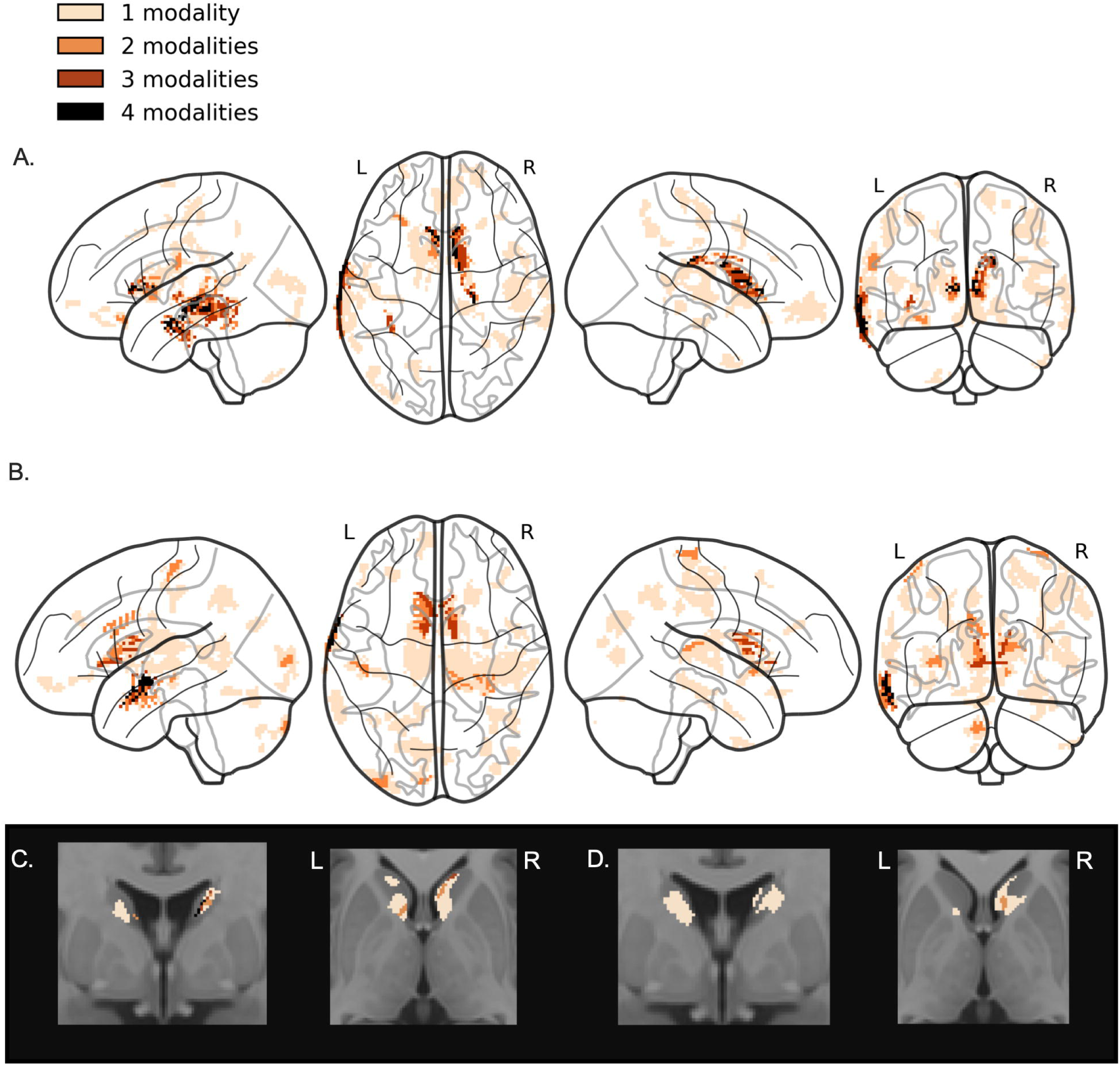

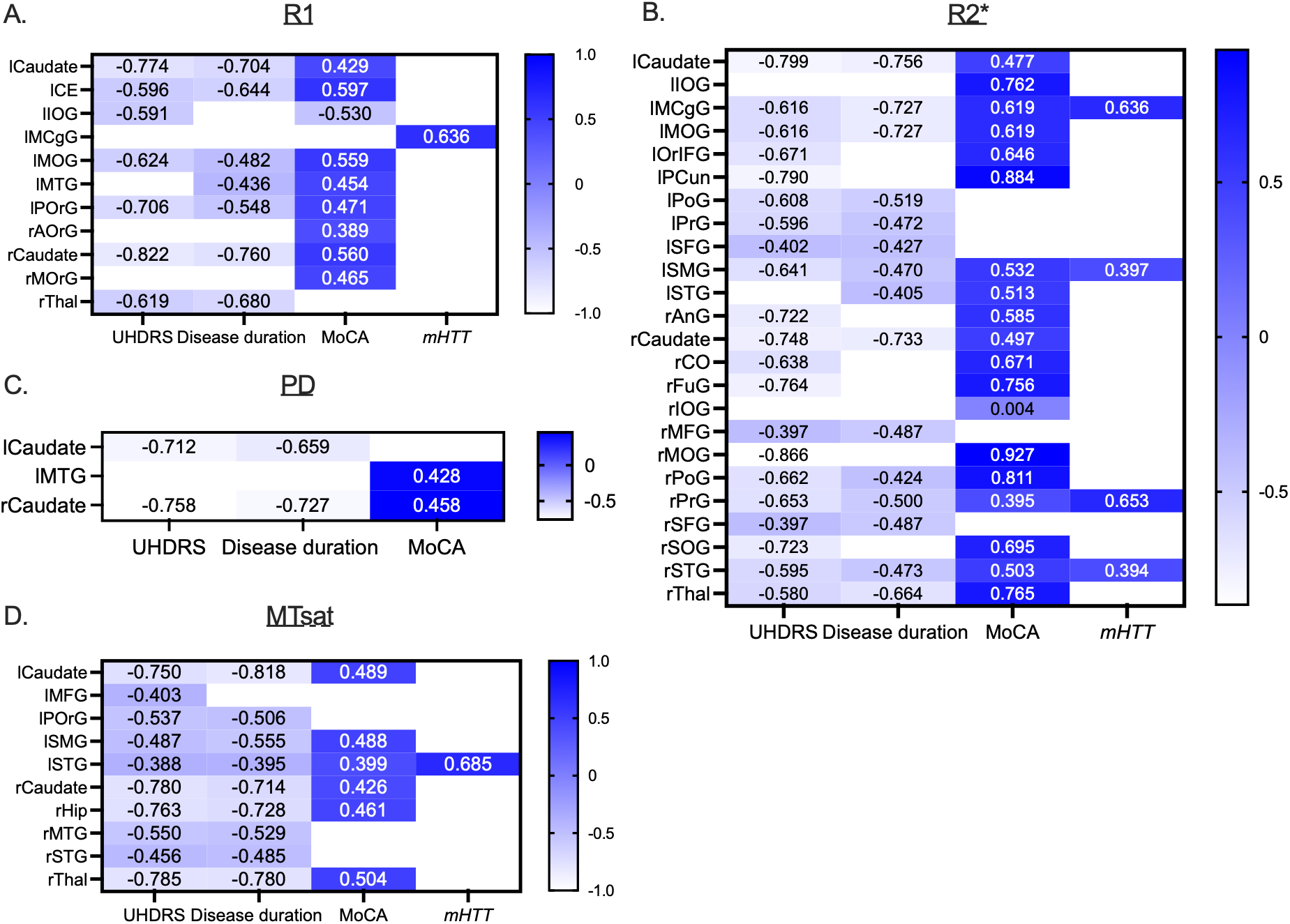

